# An Interpretable Multimodal CT-Clinical Model for Predicting 90-Day Functional Outcome After Acute Ischemic Stroke

**DOI:** 10.64898/2026.01.09.25343245

**Authors:** Wenyue Mao, Yuxiang Dai, Hang Yu, Hao Wang, Ming Yang, Wenxing Fang, Xiao Chang, Shuhan Jin, Yunman Xia, Lixiang Huang, Shuang Xia, Yuhua Fan, Weibo Chen, Zhang Shi, Chengyan Wang, He Wang

## Abstract

**BACKGROUND:** A ccurate and individualized prediction of functional outcome after acute ischemic stroke (AIS) remains challenging. Conventional prognostic markers such as baseline National Institutes of Health Stroke Scale (NIHSS) score and age show substantial inter-patient variability and do not capture the spatial heterogeneity of tissue injury and hemodynamic disturbance. Although multimodal CT imaging is routinely acquired in acute stroke care, its integration into interpretable and generalizable outcome prediction frameworks remains limited.

**METHODS:** In this multicenter retrospective study of 720 AIS patients from five institutions, we developed an interpretable multimodal prognostic model integrating non-contrast CT, CT angiography, CT perfusion, and clinical variables to predict 90-day functional outcome (mRS). The model was trained in a derivation cohort and externally validated in four independent cohorts. Feature attribution methods, including SHapley Additive exPlanations (SHAP) and Gradient-weighted Class Activation Mapping (Grad-CAM), were used to identify key predictors and derive a spatially resolved risk representation by integrating NIHSS, Tmax, and age into a composite biomarker (C-SHAP), enabling region-specific prognostic interpretation.

**RESULTS:** Among 720 patients with AIS, our model achieved superior and stable performance across the derivation and four external validation cohorts, with AUCs ranging from 0.72 to 0.82, consistently outperforming clinical-only, imaging-only, and conventional multimodal models. SHAP analysis identified baseline NIHSS, Tmax, and age as the most influential predictors, with patient-specific contributions varying by clinical context. Spatial attribution using Grad-CAM localized outcome-relevant information to functionally meaningful brain regions. The derived Combined SHAP (C-SHAP) biomarker captured regionally coherent prognostic risk patterns and showed stronger associations with 90-day mRS than lesion burden alone. Integrating C-SHAP with ischemic lesion distribution enabled more individualized and informative outcome assessment.

**CONCLUSIONS:** Integration of multimodal CT imaging with clinical variables enables accurate, generalizable, and interpretable prediction of 90-day functional outcome after acute ischemic stroke. The proposed spatially resolved risk representation extends beyond lesion-based assessment and supports individualized prognostic evaluation in clinical practice.

## Introduction

Acute ischemic stroke (AIS) is a particularly prevalent and life-threatening disease^1–3^. Commonly employed treatment options for AIS encompass intravenous administration of alteplase for thrombolysis and mechanical thrombectomy^4,5^. Accurate prediction of prognosis is very important in AIS treatment^5–8^. Clinical data have been applied in outcome prediction. For instance, the outcome of AIS patients is commonly evaluated by the modified Rankin Scale (mRS)^9^. In addition, demographic data including age and premorbid state indicators such as baseline National Institute of Health Stroke Scale (NIHSS) are important predictors of the 90-day mRS in cerebral infarction^10–12^. Meanwhile, multimodal CT, which includes non-contrast CT, CT angiography (CTA) and CT perfusion (CTP), provides complementary information on tissue status, vascular occlusion and hemodynamic compromise^13–17^. Notably, several clinical trials examining AIS prognosis, e.g., the Acute Stroke Registry and Analysis of Lausanne (ASTRAL) and DRAGON studies, recommended the joint use of clinical data and image-based indicators for prognosis^18–20^. However, these trials reported a relatively low level of predictive accuracy. Furthermore, the weights assigned to each indicator were nearly constant, resulting in a lack of personalization.

The emergence of predictive models has enabled the development of tools capable of providing personalized outcome prediction. Deep learning models are capable of processing larger-scale data and extracting additional features from multidimensional and multimodal data^21–23^. Previous studies have demonstrated that incorporating clinical factors and images improved outcome prediction in the clinical decision-making processes for AIS^24^. Applying multiple modalities, e.g., clinical factors and original images, allowed for the extraction of a more comprehensive set of data, ultimately leading to improved modeling. Several studies have combined imaging-derived features and clinical data for 90-day mRS prediction and reported prognostic accuracy compared with only using unimodal data^23,25,26^. In addition, quite a few studies have explored the impact of multimodal features on 90-day mRS prediction for thrombolysis and thrombectomy treatment, with promising results^25^. However, prior multimodal prognostic models exhibit two major limitations. First, medical imaging data are often aggressively compressed into low-dimensional representations, which can obscure or discard clinically relevant high-dimensional spatial information. Second, many such models operate as “black boxes,” offering limited insight into the mechanisms and reasoning underlying their predictions, thereby hindering clinical trust and adoption. For prognostic modeling to be clinically actionable, it must not only achieve robust predictive performance but also provide interpretable, medically meaningful explanations that align with clinical reasoning.

To address these limitations, we developed a multimodal prognostic model that integrates CT imaging and clinical data to predict 90-day mRS outcome after acute ischemic stroke. This multicenter study included a total of 720 patients from five centers, comprising one derivation cohort and four independent external validation cohorts. To enhance clinical interpretability, feature attribution methods, including SHapley Additive exPlanations (SHAP)^27^ and Gradient-weighted Class Activation Mapping (Grad-CAM)^28^, were applied to examine the consistency of model-derived predictors with established clinical and imaging knowledge. In addition, interactions between clinical variables and imaging features were evaluated at a sub–brain-region level. Based on these analyses, we derived a composite biomarker, Combined SHAP (C-SHAP), by integrating information related to key prognostic indicators, including baseline National Institutes of Health Stroke Scale (NIHSS) score, Tmax, and age. By localizing outcome-relevant information to functionally meaningful brain regions and sensitively reflecting mRS progression, C-SHAP provides a spatial representation of prognostic risk that complements conventional lesion-based assessment and may improve individualized outcome prediction.

## Methods

### Study Design and Population

This retrospective multicenter study included 720 patients with AIS between July 2018 and June 2024 (**Figure 1A**). The patient flow chart was exhibited in **Figure S1**. All patients underwent standardized multimodal CT imaging within 24 hours of symptom onset. One derivation cohort and four external validation cohorts were assembled from participating centers. The study received ethical approval from the Institutional Review Board of Zhongshan Hospital, Fudan University (No. B2022-589). All centers ensured compliance with current ethical guidelines, and informed consent was obtained for all participants. Detailed clinical characteristics of each dataset were provided in **Table 1**.

**Figure 1.**
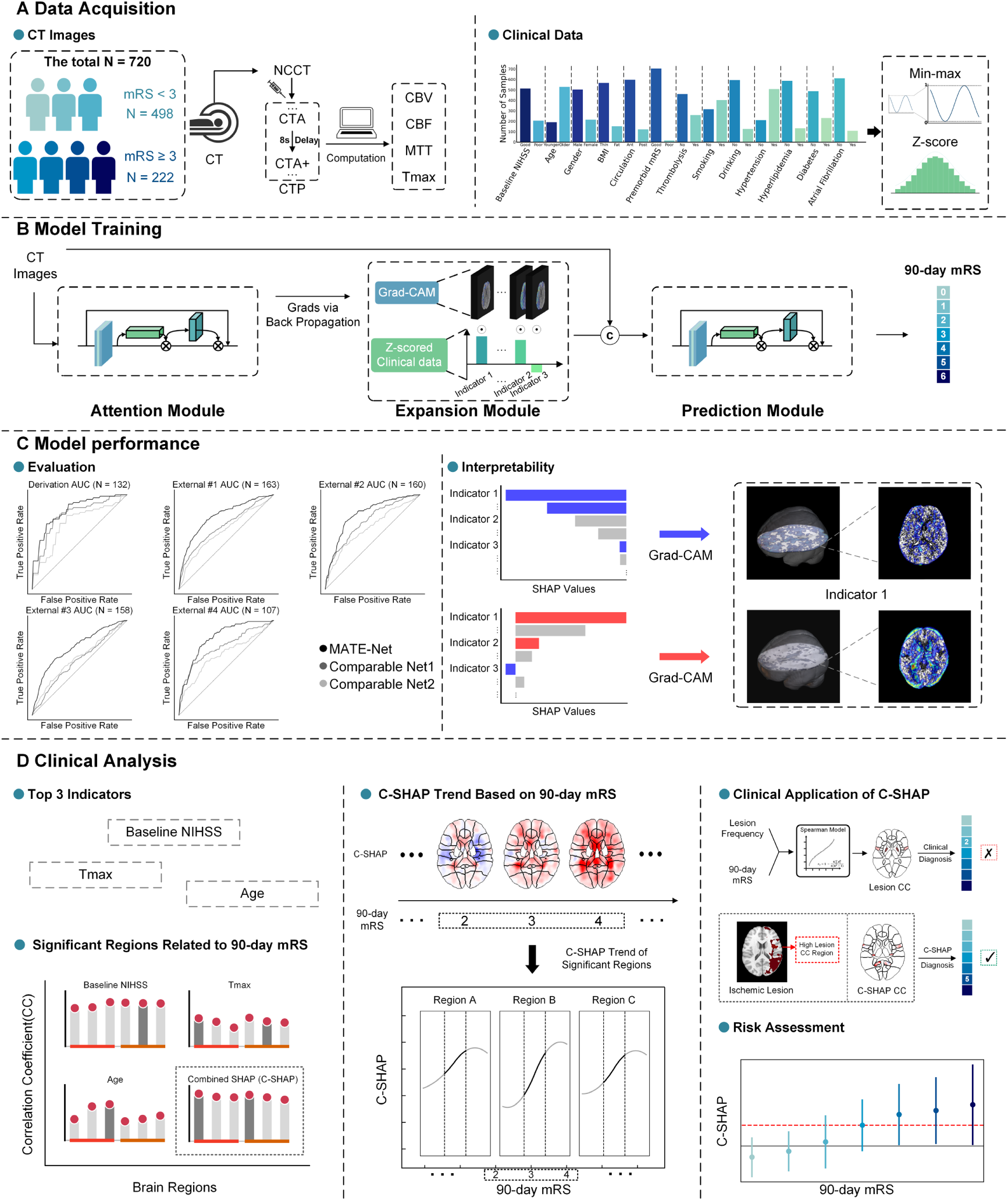
Overall framework of the study. Clinical and imaging data were collected from 720 AIS patients in five medical centers, including CT scans, demographic information, medical history, premorbid state, and 90-day mRS. In the training phase, the input indexes included non-contrast CT (NCCT), CT angiography (CTA), post-angiography CT (CTA+, acquired 8 seconds after CTA scans), cerebral blood volume (CBV), cerebral blood flow (CBF), mean transit time (MTT) and time-to-maximum (Tmax) to fit each clinical parameter in Attention Module and generate Gradient-weighted Class Activation Mappings (Grad-CAMs) in Expansion Module as clinical features. The combination of both modalities occurred at the input of Prediction Module. The clinical Grad-CAM values and CT images were concatenated and input into the module to predict the prognostic outcome of 90-day mRS. In the evaluation phase, the model’s performance was assessed primarily based on area under the receiver operating characteristic curve (AUC) and other metrics. The interpretability of the model and data was analyzed using SHapley Additive exPlanations (SHAP) values and Grad-CAMs. The spatial distribution map of SHAP values was calculated. For clinical analysis, the 3 top multimodal indicators were examined, i.e., baseline NIHSS, Tmax and age. And their interaction on brain regions were also determined. Therefore, we proposed a biomarker, i.e., Combined SHAP (C-SHAP) value. The interaction of C-SHAP on brain regions and the corresponding correlation were analyzed, and the results showed risk assessment in 90-day mRS prediction and proposed that combining ischemic lesion and C-SHAP demonstrated superior accuracy.

**Table 1.**
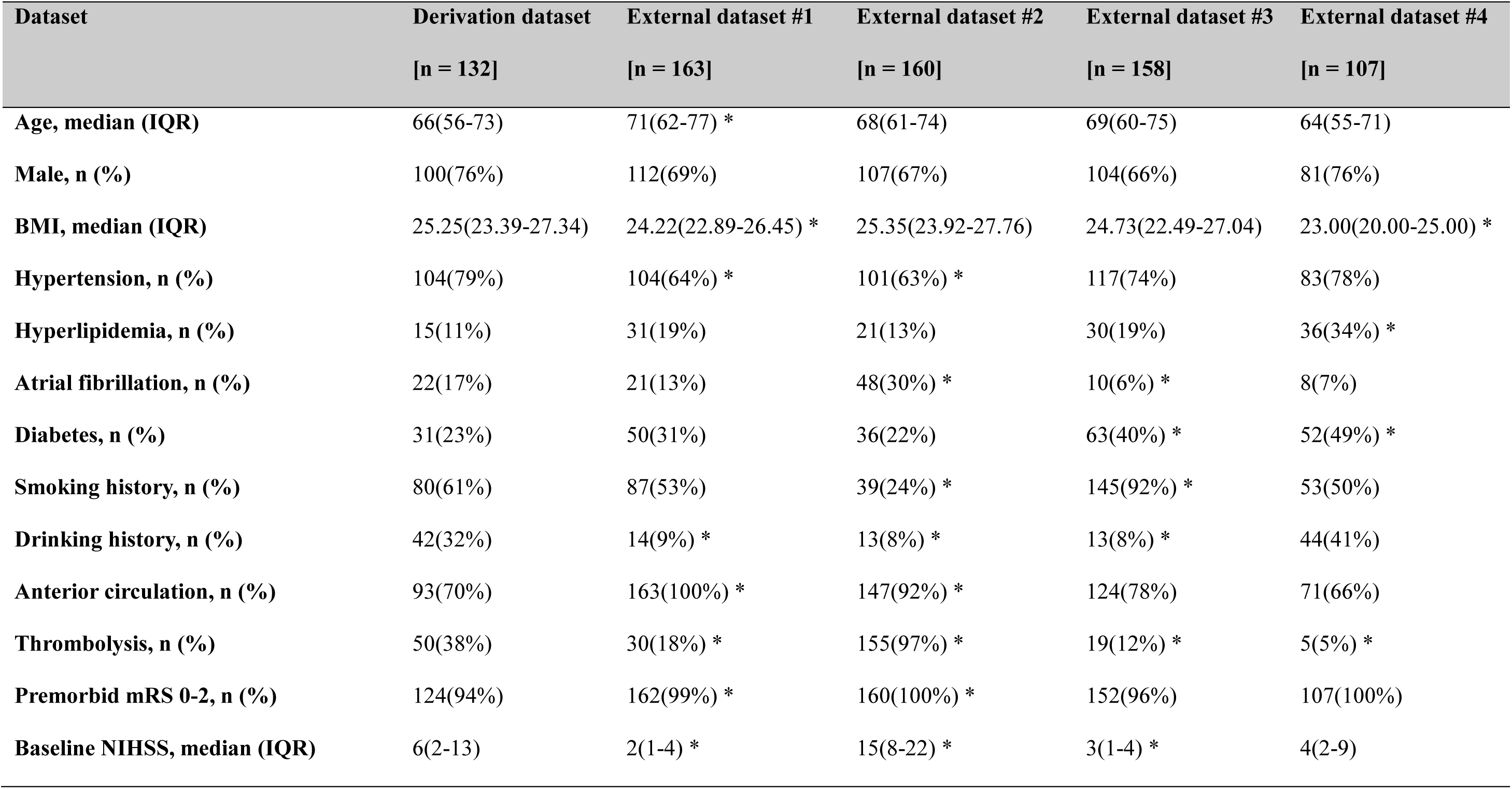
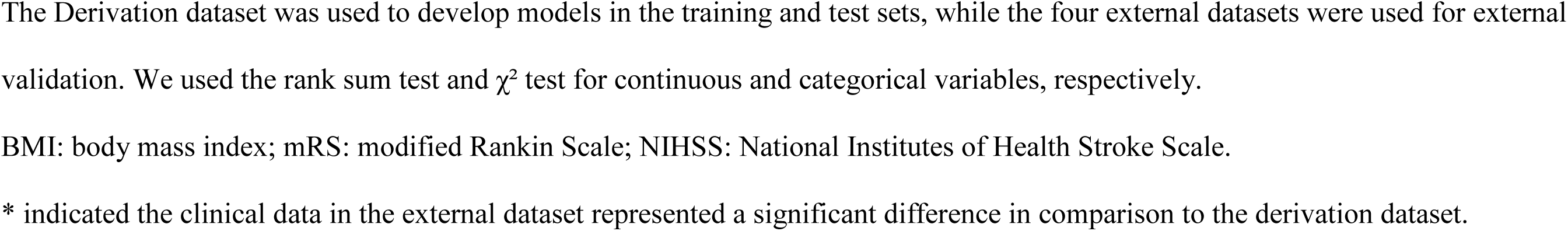
Study population and characteristics.

| Dataset | Derivation dataset | External dataset #1 | External dataset #2 | External dataset #3 | External dataset #4 |
| --- | --- | --- | --- | --- | --- |
|  | [n = 132] | [n = 163] | [n = 160] | [n = 158] | [n = 107] |
| Age, median (IQR) | 66(56-73) | 71(62-77) * | 68(61-74) | 69(60-75) | 64(55-71) |
| Male, n (%) | 100(76%) | 112(69%) | 107(67%) | 104(66%) | 81(76%) |
| BMI, median (IQR) | 25.25(23.39-27.34) | 24.22(22.89-26.45) * | 25.35(23.92-27.76) | 24.73(22.49-27.04) | 23.00(20.00-25.00) * |
| Hypertension, n (%) | 104(79%) | 104(64%) * | 101(63%) * | 117(74%) | 83(78%) |
| Hyperlipidemia, n (%) | 15(11%) | 31(19%) | 21(13%) | 30(19%) | 36(34%) * |
| Atrial fibrillation, n (%) | 22(17%) | 21(13%) | 48(30%) * | 10(6%) * | 8(7%) |
| Diabetes, n (%) | 31(23%) | 50(31%) | 36(22%) | 63(40%) * | 52(49%) * |
| Smoking history, n (%) | 80(61%) | 87(53%) | 39(24%) * | 145(92%) * | 53(50%) |
| Drinking history, n (%) | 42(32%) | 14(9%) * | 13(8%) * | 13(8%) * | 44(41%) |
| Anterior circulation, n (%) | 93(70%) | 163(100%) * | 147(92%) * | 124(78%) | 71(66%) |
| Thrombolysis, n (%) | 50(38%) | 30(18%) * | 155(97%) * | 19(12%) * | 5(5%) * |
| Premorbid mRS 0-2, n (%) | 124(94%) | 162(99%) * | 160(100%) * | 152(96%) | 107(100%) |
| Baseline NIHSS, median (IQR) | 6(2-13) | 2(1-4) * | 15(8-22) * | 3(1-4) * | 4(2-9) |

- 1 The Derivation dataset was used to develop models in the training and test sets, while the four external datasets were used for external - 2 validation. We used the rank sum test and $\chi^2$ test for continuous and categorical variables, respectively. BMI: body mass index; mRS: modified Rankin Scale; NIHSS: National Institutes of Health Stroke Scale. \* indicated the clinical data in the external dataset represented a significant difference in comparison to the derivation dataset.

Inclusion criteria were a) AIS participants with occlusions at intracranial large arteries confirmed by CT angiography (CTA) or magnetic resonance angiography (MRA), including anterior cerebral artery, middle cerebral artery, posterior cerebral artery, internal carotid artery, basilar artery and vertebral artery; b) patients receiving thrombolysis treatment (within 4.5 hours of stroke onset) and/or medical treatment (dual-antiplatelet therapy); c) patients undergoing CT examinations including non-contrast CT (NCCT), CT perfusion (CTP) on admission; d) all patients followed up after treatment and mRS score collected at 90 days.

Patients were excluded if they had: a) intracranial hemorrhage; b) thrombectomy or other surgical treatment; c) lack of the follow-up data including mRS at 90 days; d) lack of CT images or more than 30% clinical indicators; e) severe motion artifacts or poor CT image quality (quality score < 4) (**Table S1**).

### Imaging Acquisition and Preprocessing

CT scans in the used datasets were performed with the 320-Row Brilliance iCT scanner (Philips Healthcare) or SOMATOM Force scanner (Siemens Medical Solutions). Scanning parameters for NCCT were: tube voltage, 120 kV; tube current, 320 mAs; slice thickness, 5 mm; field-of-view (FOV), 30 × 30 × 16 cm^3^ covering the whole intracranial range; in-plane resolution, 0.5 × 0.5 mm^2^. Scanning parameters for CTP were: tube voltage, 80 kV; tube current, 190 mAs; FOV and spatial resolution as described for NCCT. Iodinated contrast (50mL) was injected at 5.0 mL/s, and 14 frames at time intervals of 4s were acquired. The dataset included NCCT, CTP-derived CTA and post-angiography CT (CTA+, acquired 8 seconds after CTA scans), along with perfusion parameter maps, including cerebral blood volume (CBV), cerebral blood flow (CBF), mean transit time (MTT) and time-to-maximum (Tmax), computed from raw CTP data. Ischemic core and penumbra labels were generated using RAPID^29^. All CT images across the five cohorts were preprocessed to a unified voxel size of 0.98 × 0.98 × 5.00 mm³, resampled to 256 × 256 × 32 for model input, and normalized to a 0-1 range. Additional preprocessing details are provided in the **Figures S2, S3**.

### Model Architecture: MATE-Net Overview

Out study proposed the Multimodal and ATtention-based Expansion Network (MATE-Net) to integrate 3D multimodal CT images with clinical indicators, which comprised three modules, i.e., Attention Module, Expansion Module and Prediction Module (**Figure 1B**).

Attention Module contained one model per clinical data. 7 CT images were concatenated and processed using the ResNet18^30^ augmented with the attention mechanism Convolutional Block Attention Module (CBAM)^31^, generating one-channel outputs scaled to match 0-1 standardized clinical data. This module improved feature interpretability and prepared for comprehensive feature generation in the subsequent Expansion Module.

In Expansion Module, Grad-CAM was applied to generate expanded features which were extracted from the middle layers in Attention Module. Each clinical data had a corresponding Grad-CAM, which was multiplied z-score standardized clinical value to produce expanded features. Therefore, the features reflected both the clinidal index and its associations with brain regions.

Prediction Module predicted 90-day mRS outcome using CT images concatenated with the expanded features, employing another CBAM-enhanced ResNet18 to improve predictive performance. Specific structures and details were found in **Figure S4** and **Table S2, S3**.

### Explainability and Biomarker (C-SHAP) Derivation

Model interpretability was assessed using SHapley Additive exPlanations (SHAP), which provides sample-level estimates of each feature’s contribution to the prediction. SHAP values were derived from the Prediction Module at both voxel and modality levels, and SHAP scalar values were obtained by averaging within predefined brain areas (**Figure 1C**).

The proposed spatial biomarker, combined-SHAP (C-SHAP), was generated by z-score normalizing and integrating SHAP maps associated with the key prognostic indicator (baseline NIHSS, Tmax, and age). C-SHAP was assessed for anatomical plausibility, its correlation with mRS progression, and its ability to delineate ischemic territories contributing to poor outcomes (**Figure 1D**).

### Training and Validation Strategy

The derivation cohort was used for 5-fold cross-validation, ensuring patient-level separation. External validation was performed independently on four datasets to assess generalizability. In the training phase, in Attention Module, all models had mean square error loss and applied a learning rate of 10^-6^. The predictive model selected Huber loss, with a learning rate of 10^-5^. All experiments were trained on 40GB NVIDIA A100 GPUs with CUDA 12.0. Python 3.8.13 was utilized for the software environment and PyTorch 1.13 was applied for framework establishment.

The performance of each model was evaluated by determining the mean and standard deviation of results across parameters. Model’s accuracy was obtained by receiver operating characteristic (ROC) curve analysis, calculating the area under the receiver operating characteristic curve (AUC). In addition, positive predictive value (PPV) and negative predictive value (NPV) were determined.

### Statistical Analysis

Continuous and categorical variables were compared using the rank-sum test and χ² test, respectively. Correlations and group differences in SHAP values, lesion frequencies, and outcomes were assessed using Spearman coefficients and two-tailed t-tests, with *p*-values corrected by the Benjamini–Hochberg FDR method. Confidence intervals for AUC were estimated assuming normality across cross-validation folds, and performance differences between MATE-Net and comparator models were evaluated using the DeLong test. Statistical significance was defined as *P* < 0.05. The missing data was imputed using the Random Forest model. Analyses were performed using MATLAB and Python.

## Results

A total of 720 patients were included in the analysis, consisting of 132 patients in the derivation cohort and 588 patients across four independent external validation cohorts. Baseline characteristics were balanced across datasets, although external cohorts demonstrated expected variability in age distribution, stroke severity, and imaging vendors (**Figure 2A**, **Table 1**).

**Figure 2.**
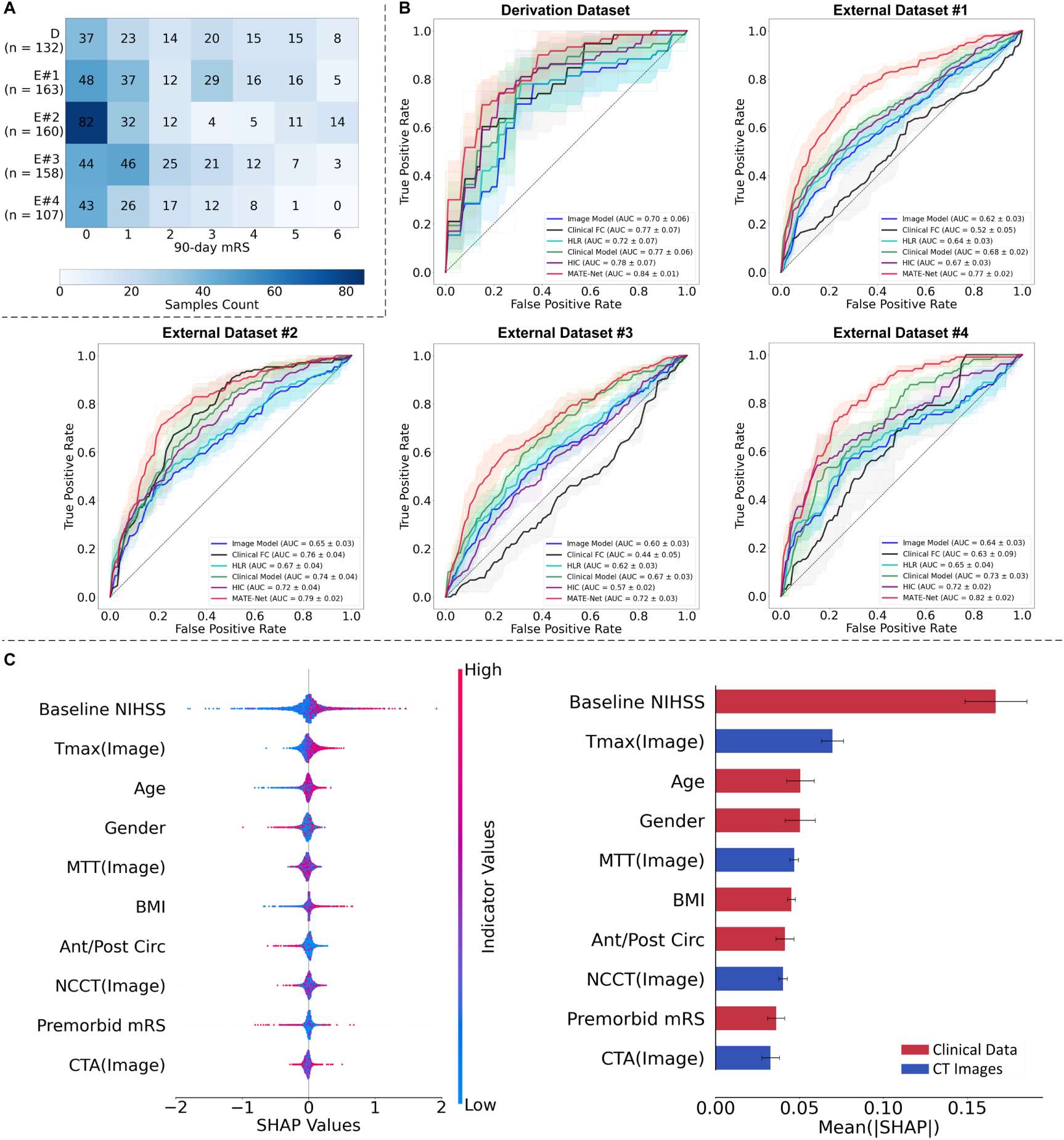
Datasets and model performances. **(A)** A tabular representation displaying the counts of 90-day mRS of the derivation and external datasets. D: Derivation dataset. E#1-E#4: External datasets #1-#4. **(B)** ROC curve analysis of the five datasets. The MATE-Net model (represented in red) outperforms other models in all datasets, with AUCs of 0.84 ± 0.01, 0.77 ± 0.02, 0.79 ± 0.02, 0.72 ± 0.03 and 0.82 ± 0.02, respectively. The other models included Image Model (only CT images), Clinical FC (Clinical Fully Connection model), HLR (Hybrid-Logistic-Regression model, a hybrid model with logistic regression), Clinical Model (only expanded clinical features), HIC (Hybrid-Image-Clinical model, another hybrid model). **(C)** The left plot presented the 10 top indicators of SHAP values generated from MATE-Net, in decreasing order of mean absolute SHAP values. The right plot showed average absolute SHAP values for the 10 top indicators in decreasing order. Indicators with higher absolute SHAP values have higher importance in AIS outcome prediction. Baseline NIHSS, Tmax and age were among the 3 top factors.

### Model performances

Across five-fold cross-validation in the derivation cohort, the proposed MATE-Net demonstrated superior predictive performance compared with all comparator models (**Table 2**). In the derivation cohort, the model achieved a mean accuracy of 0.80 ± 0.03 (95% confidence interval, CI:0.77, 0.84) and an AUC of 0.84 ± 0.01 (95% CI: 0.82,0.85).

**Table 2.**
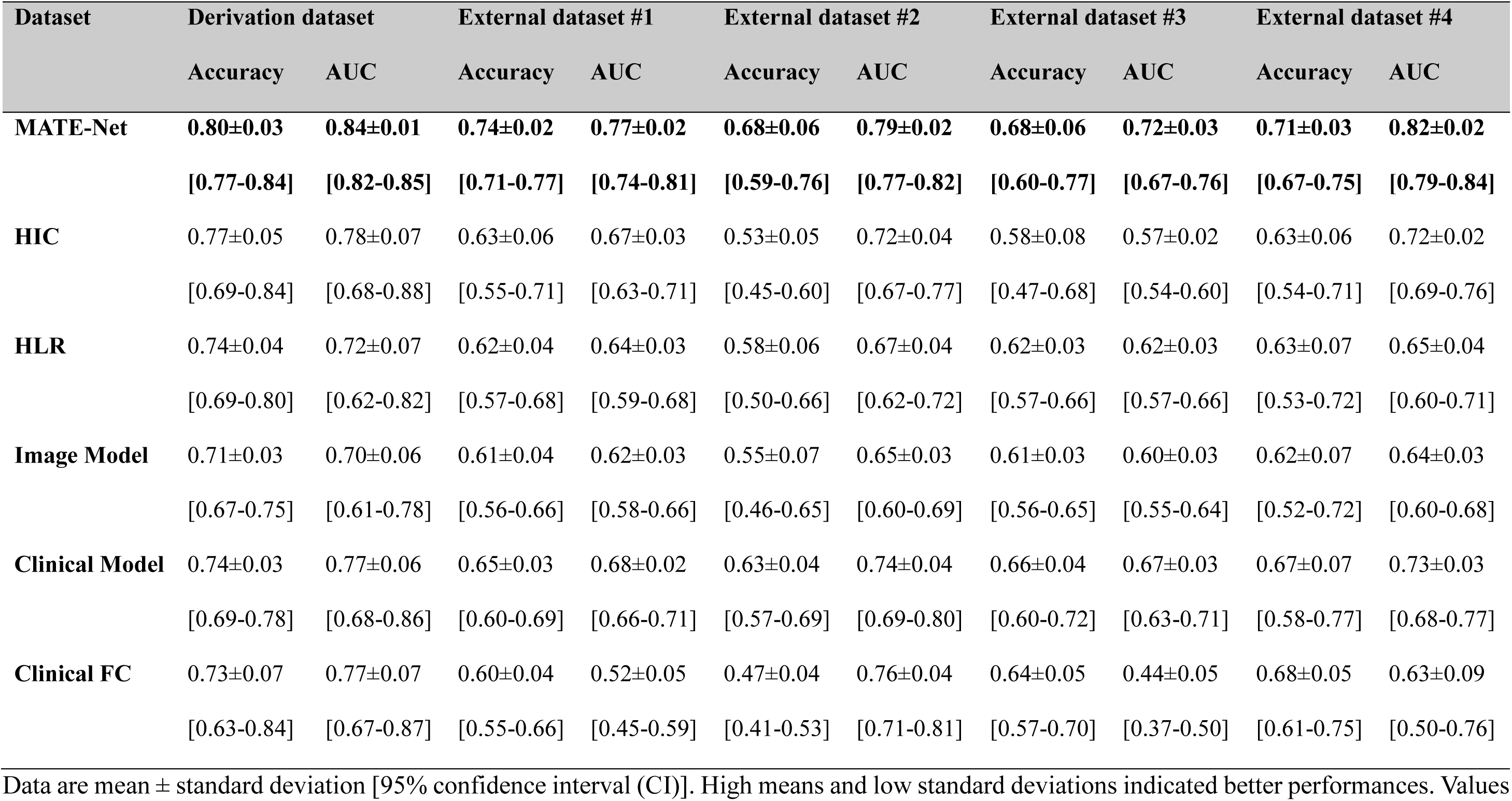
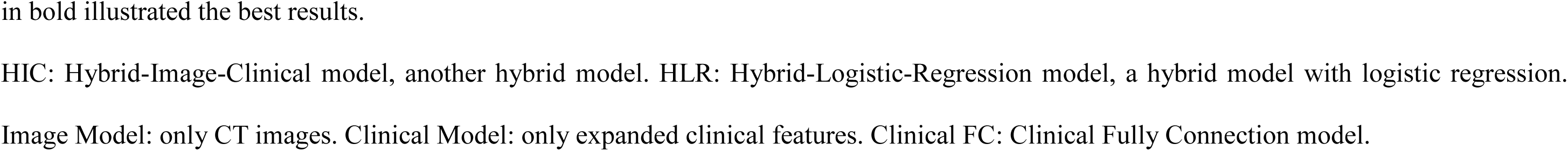
Model Performances.

| Dataset | Derivation dataset |  | External dataset #1 |  | External dataset #2 |  | External dataset #3 |  | External dataset #4 |  |
| --- | --- | --- | --- | --- | --- | --- | --- | --- | --- | --- |
|  | Accuracy | AUC | Accuracy | AUC | Accuracy | AUC | Accuracy | AUC | Accuracy | AUC |
| **MATE-Net** | **0.80±0.03** | **0.84±0.01** | **0.74±0.02** | **0.77±0.02** | **0.68±0.06** | **0.79±0.02** | **0.68±0.06** | **0.72±0.03** | **0.71±0.03** | **0.82±0.02** |
|  | [0.77-0.84] | [0.82-0.85] | [0.71-0.77] | [0.74-0.81] | [0.59-0.76] | [0.77-0.82] | [0.60-0.77] | [0.67-0.76] | [0.67-0.75] | [0.79-0.84] |
| **HIC** | 0.77±0.05 | 0.78±0.07 | 0.63±0.06 | 0.67±0.03 | 0.53±0.05 | 0.72±0.04 | 0.58±0.08 | 0.57±0.02 | 0.63±0.06 | 0.72±0.02 |
|  | [0.69-0.84] | [0.68-0.88] | [0.55-0.71] | [0.63-0.71] | [0.45-0.60] | [0.67-0.77] | [0.47-0.68] | [0.54-0.60] | [0.54-0.71] | [0.69-0.76] |
| **HLR** | 0.74±0.04 | 0.72±0.07 | 0.62±0.04 | 0.64±0.03 | 0.58±0.06 | 0.67±0.04 | 0.62±0.03 | 0.62±0.03 | 0.63±0.07 | 0.65±0.04 |
|  | [0.69-0.80] | [0.62-0.82] | [0.57-0.68] | [0.59-0.68] | [0.50-0.66] | [0.62-0.72] | [0.57-0.66] | [0.57-0.66] | [0.53-0.72] | [0.60-0.71] |
| **Image Model** | 0.71±0.03 | 0.70±0.06 | 0.61±0.04 | 0.62±0.03 | 0.55±0.07 | 0.65±0.03 | 0.61±0.03 | 0.60±0.03 | 0.62±0.07 | 0.64±0.03 |
|  | [0.67-0.75] | [0.61-0.78] | [0.56-0.66] | [0.58-0.66] | [0.46-0.65] | [0.60-0.69] | [0.56-0.65] | [0.55-0.64] | [0.52-0.72] | [0.60-0.68] |
| **Clinical Model** | 0.74±0.03 | 0.77±0.06 | 0.65±0.03 | 0.68±0.02 | 0.63±0.04 | 0.74±0.04 | 0.66±0.04 | 0.67±0.03 | 0.67±0.07 | 0.73±0.03 |
|  | [0.69-0.78] | [0.68-0.86] | [0.60-0.69] | [0.66-0.71] | [0.57-0.69] | [0.69-0.80] | [0.60-0.72] | [0.63-0.71] | [0.58-0.77] | [0.68-0.77] |
| **Clinical FC** | 0.73±0.07 | 0.77±0.07 | 0.60±0.04 | 0.52±0.05 | 0.47±0.04 | 0.76±0.04 | 0.64±0.05 | 0.44±0.05 | 0.68±0.05 | 0.63±0.09 |
|  | [0.63-0.84] | [0.67-0.87] | [0.55-0.66] | [0.45-0.59] | [0.41-0.53] | [0.71-0.81] | [0.57-0.70] | [0.37-0.50] | [0.61-0.75] | [0.50-0.76] |

2 Data are mean ± standard deviation [95% confidence interval (CI)]. High means and low standard deviations indicated better performances. Values
- 1 **in bold** illustrated the best results. HIC: Hybrid-Image-Clinical model, another hybrid model. HLR: Hybrid-Logistic-Regression model, a hybrid model with logistic regression. Image Model: only CT images. Clinical Model: only expanded clinical features. Clinical FC: Clinical Fully Connection model.

Importantly, model performance remained stable across all four external validation cohorts. Accuracy ranged from 0.68 to 0.74, and AUC values ranged from 0.72 to 0.82, indicating robust generalizability across heterogeneous patient populations and imaging environments. Positive and negative predictive values were consistently higher than those of comparator approaches (**Table S4**).

In the derivation cohort (**Figure 2B**), MATE-Net (AUC = 0.84) surpassed all comparative models, including HIC (Hybrid-Image-Clinical model, another hybrid model; AUC = 0.78), followed by Clinical Model (only input with expanded clinical features, AUC = 0.77), Clinical FC (Clinical Fully Connection model, only input with clinical data; AUC = 0.77), HLR (Hybrid-Logistic-Regression model, another hybrid model with logistic regression; AUC = 0.72), and Image Model (only input with CT images, AUC = 0.70). Similar trends were observed across external datasets, where MATE-Net consistently exhibited the highest average AUCs. In contrast, Clinical FC showed marked instability across sites. DeLong tests confirmed that MATE-Net significantly outperformed all comparator models (**Table S5**).

Overall, these results demonstrated that the multimodal design of MATE-Net provided clear and consistent performance gains over unimodal and feature compression multimodal approaches, with expanded feature representations contributing substantially to improved prognostic accuracy.

### Indicator contributions for AIS outcome prediction

SHAP analysis was applied and showed that baseline NIHSS (mean absolute SHAP, SHAP = 0.17, standard deviation, std = 0.02), Tmax (SHAP = 0.07, std = 0.01) and age (SHAP = 0.05, std = 0.01) were the 3 most influential predictors in MATE-Net (**Figure 2C**), consistent with the leading indicators identified in unimodal image and clinical models (**Figure S5**). Ablation experiments further confirmed their importance (**Table S6**). When both baseline NIHSS and Tmax were excluded, the performance was worse(AUC from 0.84 to 0.76).

Case-level analyses demonstrated individualized contributions of clinical and imaging factors to outcome prediction. Among patients with comparable age and lesion extent, higher baseline NIHSS scores were associated with worse functional outcomes, reflected by larger SHAP contributions of NIHSS (**Figure 3A**). When baseline neurological deficits and lesion burden were mild, age became the dominant contributor, indicating increased vulnerability to poor recovery in older patients despite limited initial injury (**Figure 3B**). In patients with intermediate baseline NIHSS scores and age, perfusion-related injury, particularly increased Tmax delay, played a more prominent prognostic role (**Figure 3C**).

**Figure 3.**
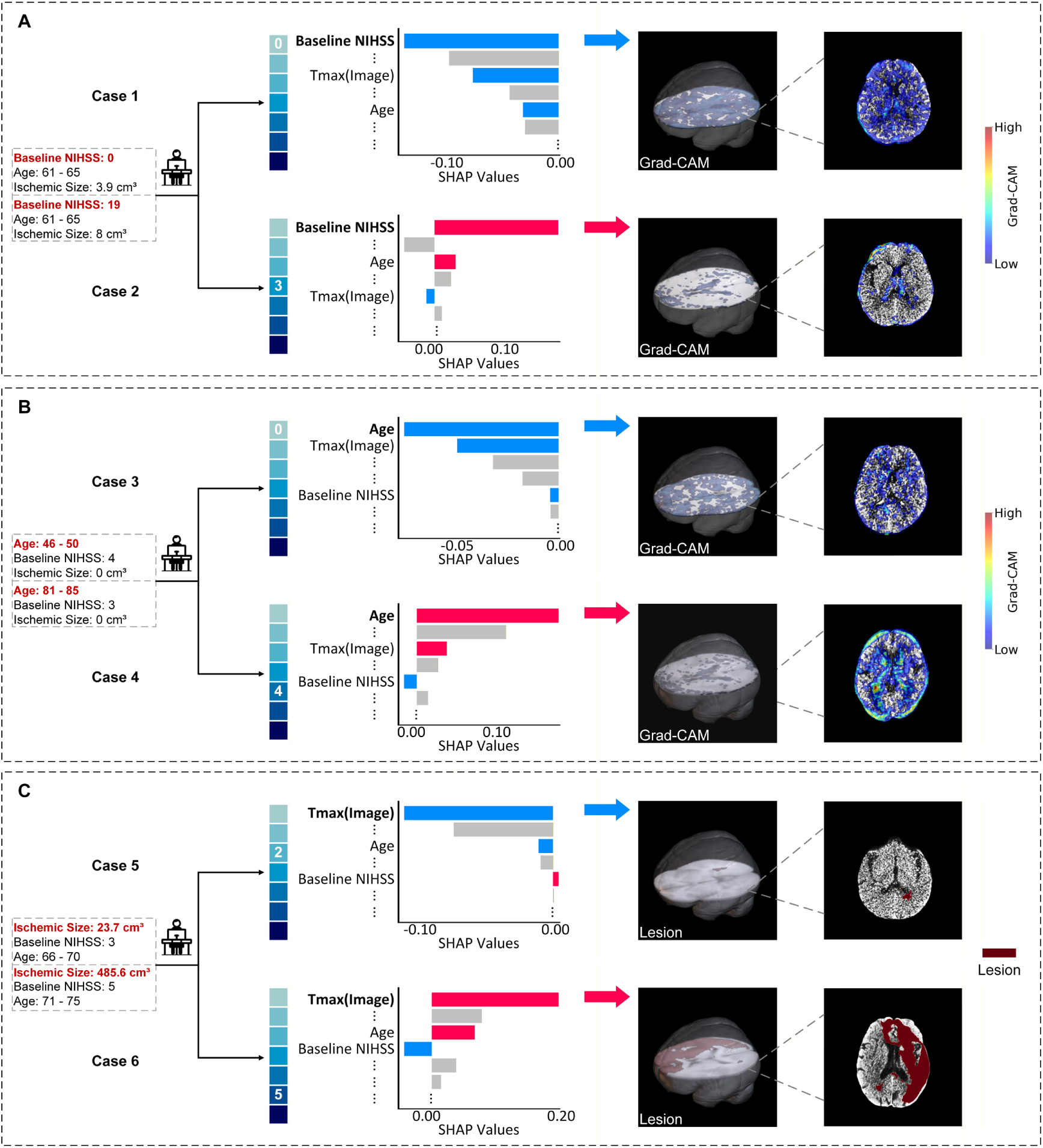
Typical cases to show the diagnostic values of top SHAP indicators for mRS prediction. Each row illustrated an individual case with corresponding SHAP values and Grad-CAMs, in which the color bar (blue and red) emphasized the most important indicator as ranked by SHAP. This ischemic lesion was assessed with Tmax > 6s. **(A)** Two cases were shown with similar lesion sizes and ages, but different baseline NIHSS values, which led to significantly different 90-day mRS scores and was effectively reflected by SHAP values. In the corresponding Grad-CAM from baseline NIHSS, the sample with a baseline NIHSS of 19 was more focused on details in the central structures. **(B)** Two samples were displayed with no lesion and the same baseline NIHSS, but distinct ages and different outcomes. When baseline NIHSS and lesion size were not notable, the older case had higher SHAP value, and the corresponding Grad-CAM focused more on ventricles. **(C)** Two samples were depicted with comparable baseline NIHSS values and ages, but completely different ischemic lesion sizes and outcomes. These groups illustrated the significant effect of Tmax with baseline NIHSS being neither high nor low.

Gradient-weighted Class Activation Mapping (Grad-CAM) was used to localize outcome-relevant brain regions associated with influential clinical variables in patients with poor outcomes. In **Figure 3A**, NIHSS-related Grad-CAM activation was concentrated within deep gray matter structures, including the thalamus and basal ganglia. In contrast, age-related Grad-CAM activation in **Figure 3B** was predominantly localized around the ventricular system. These findings highlight patient-specific interactions between clinical severity, perfusion abnormalities, and regional brain vulnerability.

### Construction and Evaluation of the proposed C-SHAP biomarker

Based on the feature importance ranking (**Figure 2C**), we further investigated how baseline NIHSS, age, and Tmax contributed to outcome prediction at the regional brain level. SHAP maps corresponding to these indicators were registered to MNI-152 space^32^ and segmented using the Hammers atlas^33–35^. Correlations between regional SHAP values within anterior circulation territories and 90-day mRS scores were calculated (**Figure 4A, Figure S6**, **Table S7**).

**Figure 4.**
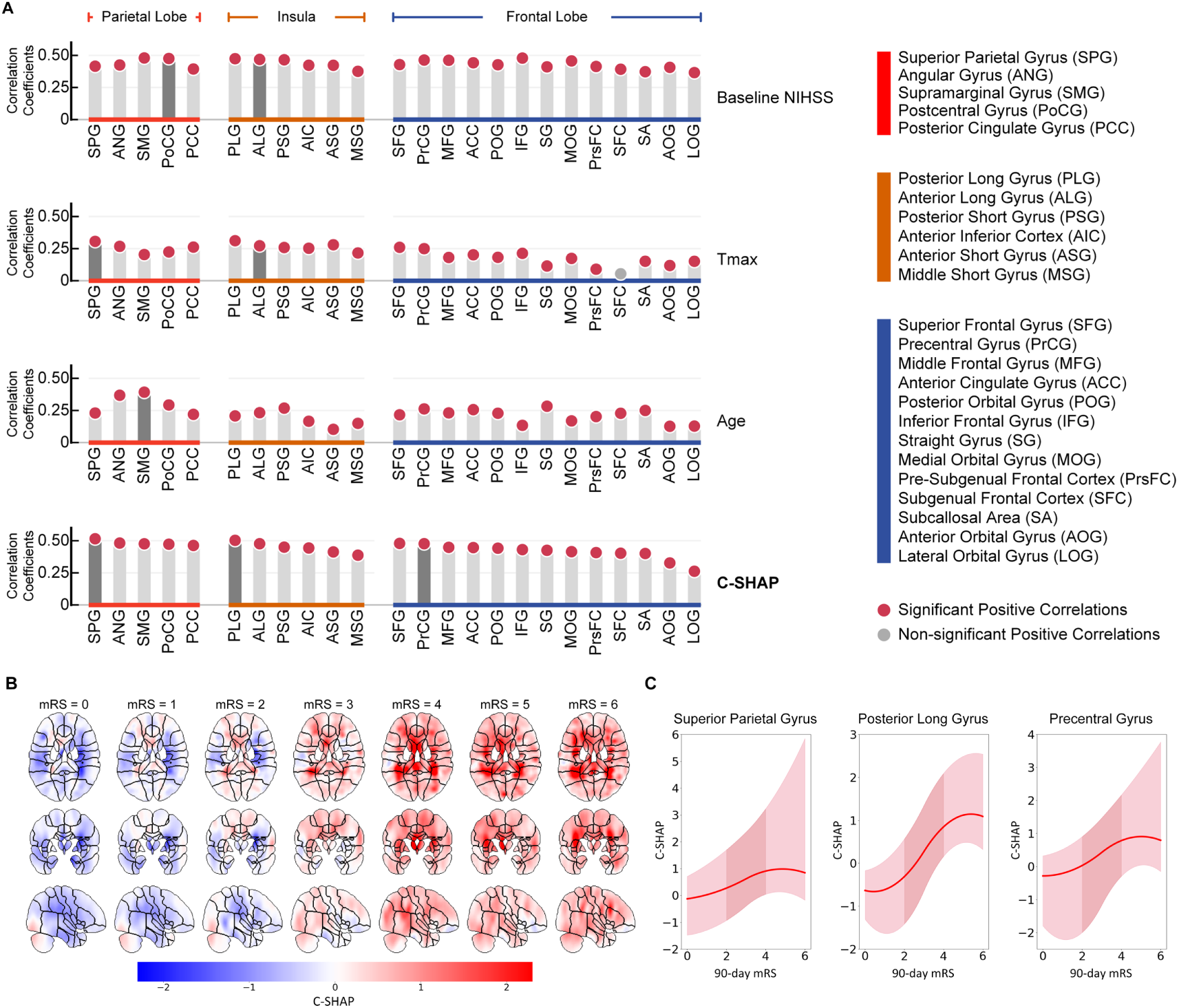
Interactions between clinical data and brain regions. **(A)** The correlation between SHAP values of anterior circulation brain regions and 90-day mRS was calculated for the 3 top indicators (baseline NIHSS, Tmax and age) and the Combined SHAP (C-SHAP), and the outcomes were exhibited in decreasing order of the sub brain region correlations of C-SHAP. Consistently correlated brain regions (parietal lobe, insula and frontal lobe), i.e., most frequently ranked among the 10 top high correlations across various datasets, were portrayed, and results on other brain regions can be found in Figure S5. **(B)** C-SHAP on brain maps for mRS = 0 to 6. **(C)** Trend of average C-SHAP regions with increasing mRS score in the consistently strong correlated regions from **(A)**, i.e., superior parietal gyrus, posterior long gyrus and precentral gyrus. Detailed correlation coefficients can be found in Table S6.

For baseline NIHSS, the postcentral gyrus (correlation coefficient across all datasets, cc = 0.471, *P* < 0.05) and the anterior long gyrus (cc = 0.464, *P* < 0.05) showed consistently strong correlations with functional outcome and were most frequently ranked among the top ten regions across datasets. For Tmax, significant correlations were observed in the superior parietal gyrus (cc = 0.302, *P* < 0.05) and the anterior long gyrus (cc = 0.268, *P* < 0.05). In contrast, age-related SHAP values were most strongly associated with regions including the supramarginal gyrus (cc = 0.388, *P* < 0.05), posterior temporal lobe (cc = 0.381, *P* < 0.05), and the middle and inferior temporal gyri (cc = 0.339, *P* < 0.05). The variability in regional associations across individual indicators highlighted the limitations of single-feature interpretability for capturing patient-specific prognostic patterns.

To integrate these complementary effects, we constructed the Combined SHAP (C-SHAP) biomarker by z-score normalizing and aggregating SHAP maps from the three dominant predictors. We then examined changes in C-SHAP across increasing levels of disability, focusing on the clinically relevant mRS range of 2 to 4 (**Figure 4B**). As functional outcome worsened, positive C-SHAP values progressively expanded within the superior parietal gyrus (mean C-SHAP: 0.27 to 0.90), posterior long gyrus (−0.31 to 0.85), and precentral gyrus (0.03 to 0.78), indicating increasing regional prognostic risk (**Figure 4C**). These findings suggested that C-SHAP captured spatially coherent, outcome-relevant patterns that may support region-specific prognostic assessment and inform individualized therapeutic decision-making.

### Integration of ischemic lesions and C-SHAP for AIS prognosis

Ischemic lesions were defined using a Tmax threshold greater than 6 seconds, and ischemic core was defined as regions with relative cerebral blood flow (rCBF) less than 30%. Although ischemic core volume showed a stronger overall correlation with 90-day mRS outcomes, reliable estimation of core lesions in the hyperacute stage remains clinically challenging. Therefore, subsequent analyses focused on ischemic lesions defined by Tmax delay.

Within the anterior circulation, the supramarginal gyrus exhibited the highest lesion frequency (**Figure 5A**); however, its correlation with functional outcome was weak (cc = 0.090). In contrast, lesions involving the precentral gyrus (cc = 0.156, *P* < 0.05) and the pallidum (cc = 0.200, *P* < 0.05) showed more consistent associations with worse functional outcomes, suggesting that involvement of these regions may carry greater prognostic significance. Detailed regional statistics are provided in **Table S8**.

**Figure 5.**
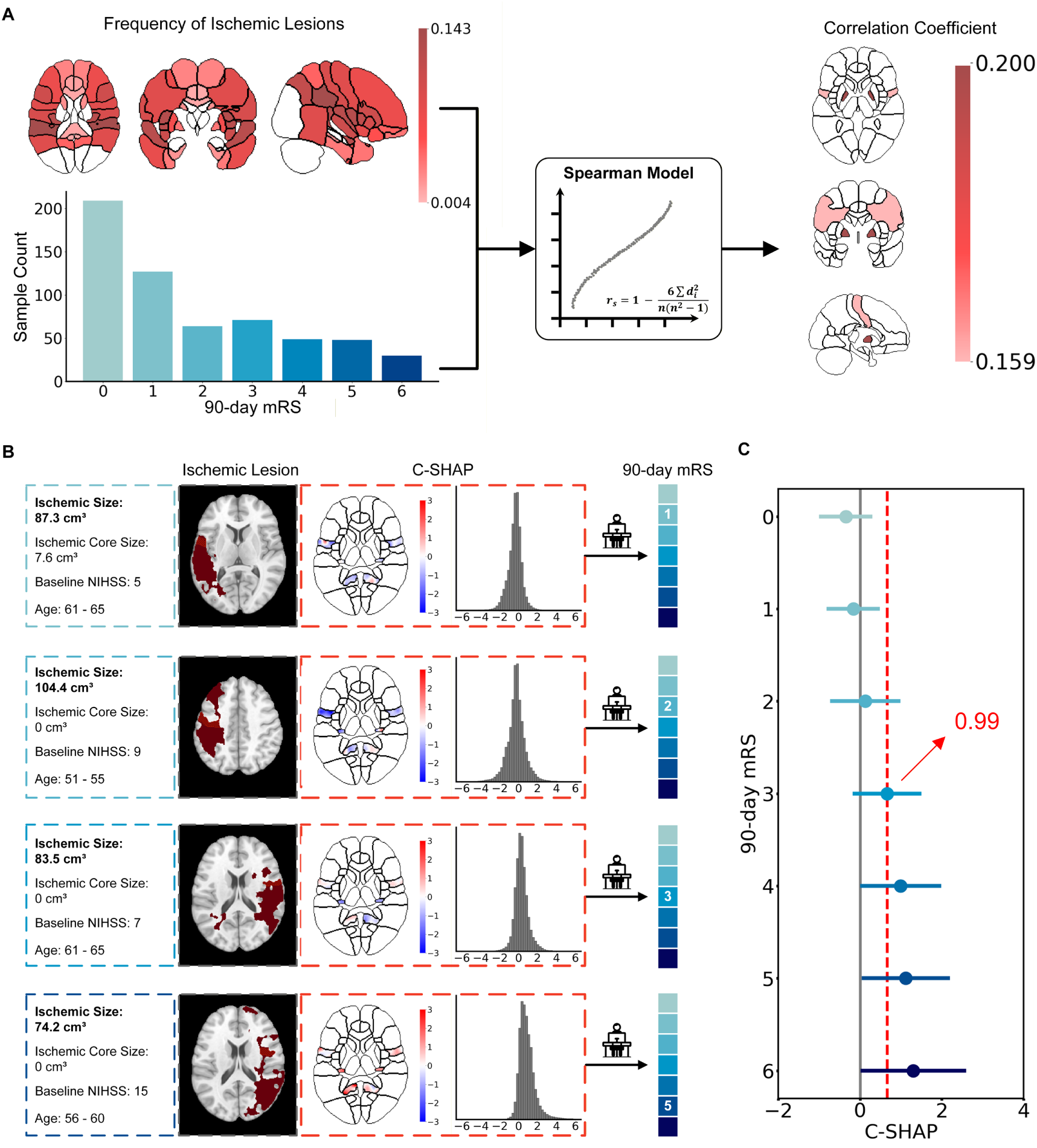
C-SHAP complement the analysis of ischemic lesions. **(A)** Average frequency of ischemic lesions in the anterior circulation. The correlation between the frequency of ischemic lesions and 90-day mRS was depicted, and precentral gyrus and pallidum showed the consistent correlations. **(B)** Four samples portrayed in increasing order of 90-day mRS. All samples had similar ages, while the ischemic sizes and baseline NIHSS values were not arranged in increasing order. The positive C-SHAP values increased significantly as 90-day mRS increased. **(C)** C-SHAP was applied for risk assessment. The trend was noticeable that median C-SHAP value became positive as 90-day mRS ranged. It was noteworthy that the median C-SHAP value was 0.99 when 90-day mRS was 3, and the higher C-SHAP indicated a poor outcome.

Because lesion distribution alone or C-SHAP alone was insufficient to fully explain outcome heterogeneity, their combined prognostic value was further evaluated (**Figure 5B**). Case-level analyses demonstrated that patients with similar lesion distributions could experience markedly different functional outcomes, particularly when lesions involved mRS-relevant regions such as the precentral gyrus, underscoring the limitations of lesion-based assessment alone. When C-SHAP was incorporated, positive values within consistently correlated regions, including the superior parietal, posterior long, and precentral gyri, progressively increased with worsening mRS.

For example, despite having a larger ischemic lesion burden, one patient with low C-SHAP values in these regions achieved a more favorable outcome compared with another patient who had smaller lesions but higher regional C-SHAP values. Furthermore, C-SHAP values within strongly correlated regions increased substantially as 90-day mRS worsened, particularly across the clinically relevant range from mRS 2 to 4 (**Figure 5C**). When regional C-SHAP values exceeded 0.99, corresponding to the median C-SHAP observed in patients with mRS = 3, this was associated with a transition toward poorer functional outcome.

Overall, integrating ischemic lesion distribution with C-SHAP provided a more informative and individualized assessment of functional prognosis than lesion-based metrics alone, supporting its potential utility in personalized mRS prediction and clinical decision-making.

## Discussion

In this study, we developed MATE-Net, a multimodal prognostic framework that integrates routinely acquired CT imaging and clinical variables to predict 90-day functional outcome after acute ischemic stroke. By preserving spatial information from imaging data and explicitly linking it to key clinical indicators, MATE-Net achieved improved predictive performance while maintaining clinical interpretability. The model consistently identified baseline NIHSS, Tmax, and age as the dominant contributors to outcome prediction across derivation and external validation cohorts.

A key methodological feature of MATE-Net is the use of Grad-CAM–based feature expansion to preserve anatomical structure and facilitate interpretable multimodal integration. Rather than relying on abstract deep features, this approach enabled localization of outcome-relevant information to specific brain regions associated with clinical severity. Grad-CAM visualizations linked baseline NIHSS to deep gray matter structures, including the thalamus and basal ganglia, consistent with their roles in consciousness, motor function, and language processing reported in prior studies^36–39^. Age-related activation patterns were predominantly localized around the ventricular system, reflecting structural vulnerability associated with aging^40,41^. These findings support the biological plausibility of the model and demonstrate how spatial information can enhance interpretability of multimodal outcome prediction.

SHAP analysis further quantified the relative importance of individual predictors and provided a foundation for construction of the C-SHAP biomarker. By integrating contributions from baseline NIHSS, Tmax, and age into a unified spatial representation, C-SHAP captured region-specific vulnerability that was not apparent from lesion extent or single indicators alone. C-SHAP values showed consistent associations with 90-day mRS outcomes and evolved progressively with increasing disability, particularly within the clinically relevant mRS range of 2 to 4. Higher C-SHAP values in functionally critical regions were associated with worse outcomes, supporting its potential role as a spatially resolved imaging biomarker for individualized prognostic assessment.

Importantly, our results highlight the limitations of lesion-based assessment alone for outcome prediction. Patients with similar ischemic lesion distributions exhibited markedly different functional outcomes, whereas integration of lesion information with C-SHAP better reflected interindividual variability. This combined approach provides a more comprehensive representation of stroke-related brain injury by accounting for both anatomical damage and patient-specific susceptibility, which may be relevant for clinical decision-making and risk stratification.

Several limitations should be acknowledged. High-quality CT imaging is required for reliable feature extraction, particularly for perfusion-derived parameters such as Tmax. Future work may explore incorporation of higher-resolution imaging modalities, including diffusion-weighted MRI, to further enhance spatial characterization of ischemic injury. In addition, the interpretability of Grad-CAM depends on the underlying network architecture, underscoring the importance of careful model design. Our study cohort primarily included patients treated with intravenous thrombolysis; inclusion of patients undergoing endovascular therapy and validation in broader stroke populations will be important for generalizability. Finally, although this study was retrospective, the proposed framework warrants evaluation in prospective clinical settings.

In conclusion, MATE-Net provides an interpretable and generalizable multimodal framework for prediction of 90-day functional outcome after acute ischemic stroke. The proposed C-SHAP biomarker offers a region-based representation of prognostic risk that integrates clinical severity, hemodynamic impairment, and patient vulnerability, outperforming lesion-based metrics and single indicators alone. These findings support the potential utility of spatially resolved imaging biomarkers for personalized outcome prediction and future clinical translation.

## Data Availability

The data that support the findings of this study are not publicly available due to ethical and privacy restrictions but are available from the corresponding author upon reasonable request.

## Acknowledgements

Author contributions:

Conceptualization: W.M., Y.D., X.C., Y.X., Z.S., C.W., He. W.

Data curation: W.M.

Formal analysis: W.M., Y.D., Z.S., C.W.

Investigation: W.M.

Methodology: W.M., Y.D., X.C., Y.X., Z.S., C.W., He. W.

Resources: M.Y., W.F., Hao. W, L.H, S.X, Y.F, He. W.

Software: W.M., M.Y., W.F., He. W.

Visualization: W.M., Y.D.

Writing — original draft: W.M., Y.D.

Writing — review & editing: W.M., Y.D., H.Y., Hao. W, S.J., Z.S., C.W.

## Data and materials availability

All data needed to evaluate the conclusions in the paper are present in the paper and/or the Supplementary Materials. Python codes are available on GitHub (https://github.com/Yunena/MATE-Net).

## Abbreviations

AIS: acute ischemic stroke
AUC: area under the receiver operating characteristics curve
BMI: body mass index
CBF: cerebral blood flow
CBV: cerebral blood volume
CI: confidence interval
C-SHAP: Combined SHAP
CTA: CT angiography
CTA+: post-angiography CT
CTP: CT perfusion
Grad-CAM: Gradient Class Activation Mapping
MATE-Net: Multimodal and ATtention-based Expansion Network
MRA: magnetic resonance angiography
mRS: modified Rankin Scale
MTT: transitmean transit time
NCCT: contrastnon-contrast CT
NIHSS: National Institutes of Health Stroke Scale
SHAP: SHapley Additive exPlanations
Tmax: time-to-maximum

