## Supplementary Material for "An Interpretable Multimodal CT-Clinical Model for Predicting 90-Day Functional Outcome After Acute Ischemic Stroke"

**Supplemental Material**

Tables S1-S8

Figures S1-S6

### Supplementary Tables

**Table S1 Scoring criteria for image quality**

| Score | Grade | Details |
|---|---|---|
| 5 | Excellent | No artifact and noise are present. The image is clear, with well-preserved details, uniform intensity, and strong contrast. |
| 4 | Good | Minor artifacts and noise are present. The image is relatively clear, with slightly uneven intensity but well-defined contrast, which does not affect most of the analysis. |
| 3 | Moderate | Quite apparent artifacts and noise are present. The image has uneven brightness, with poor contrast areas in field of view, resulting in difficult details recognition. |
| 2 | Poor | Severe artifact and noise are present. The image is blurry with low contrast and unrecognizable structures. |
| 1 | Unavailable | The image is severely blurred, with no discernible structures, making analysis impossible. |

**Table S2 Architecture of Attention Module and Prediction Module**

| | Attention Module | Prediction Module |
|---|---|---|
| conv0 | 7×7×7, 64 | CBAM (20) |
| layer1 | $\begin{bmatrix} 3\times 3\times 3, 64 \\ 3\times 3\times 3, 64 \\ \text{CBAM}(64) \end{bmatrix}\times 2$ | $\begin{bmatrix} 3\times 3\times 3, 64 \\ 3\times 3\times 3, 64 \\ \text{CBAM}(64) \end{bmatrix}\times 2$ |
| layer2 | $\begin{bmatrix} 3\times 3\times 3, 128 \\ 3\times 3\times 3, 128 \\ \text{CBAM}(128) \end{bmatrix}\times 2$ | $\begin{bmatrix} 3\times 3\times 3, 128 \\ 3\times 3\times 3, 128 \\ \text{CBAM}(128) \end{bmatrix}\times 2$ |
| layer3 | $\begin{bmatrix} 3\times 3\times 3, 256 \\ 3\times 3\times 3, 256 \\ \text{CBAM}(256) \end{bmatrix}\times 2$ | $\begin{bmatrix} 3\times 3\times 3, 256 \\ 3\times 3\times 3, 256 \\ \text{CBAM}(256) \end{bmatrix}\times 2$ |
| layer4 | $\begin{bmatrix} 3\times 3\times 3, 512 \\ 3\times 3\times 3, 512 \\ \text{CBAM}(512) \end{bmatrix}\times 2$ | $\begin{bmatrix} 3\times 3\times 3, 512 \\ 3\times 3\times 3, 512 \\ \text{CBAM}(512) \end{bmatrix}\times 2$ |
| fully-connected layers | 512,1 | 512,1 |

For conv0 and layers, the first part represents the kernel size of convolutional networks, and the second part expresses the output size. For fc, the first part shows the input size while the second is the output size.

**Table S3 Architecture of Convolutional Block Attention Module (CBAM)**

| | **CBAM (INPUT_SIZE)** |
|---|---|
| Channel Attention Module | 3×3×3, 1 |
| | 3×3×3, INPUT_SIZE |
| Spatial Attention Module | 7×7×7, 1 |

The first part represents the kernel size of convolutional networks, and the second expresses the output size.

**Table S4 Models Performances.**

| Models | MATE-Net | HIC | HLR | Image Model | Clinical Model | Clinical FC |
|---|---|---|---|---|---|---|
| **Derivation Dataset** | | | | | | |
| Accuracy | **0.80±0.03** | 0.77±0.05 | 0.74±0.04 | 0.71±0.03 | 0.74±0.03 | 0.73±0.07 |
| AUC | **0.84±0.01** | 0.78±0.07 | 0.72±0.07 | 0.70±0.06 | 0.77±0.06 | 0.77±0.07 |
| PPV | 0.79±0.06 | 0.73±0.07 | 0.73±0.07 | 0.71±0.06 | 0.72±0.04 | 0.71±0.08 |
| NPV | **0.82±0.04** | 0.82±0.10 | 0.75±0.03 | 0.72±0.02 | 0.75±0.04 | 0.75±0.07 |
| **External Dataset #1** | | | | | | |
| Accuracy | **0.74±0.02** | 0.63±0.06 | 0.62±0.04 | 0.61±0.04 | 0.65±0.03 | 0.60±0.04 |
| AUC | **0.77±0.02** | 0.67±0.03 | 0.64±0.03 | 0.62±0.03 | 0.68±0.01 | 0.52±0.05 |
| PPV | **0.72±0.09** | 0.56±0.08 | 0.54±0.05 | 0.53±0.05 | 0.59±0.04 | 0.57±0.20 |
| NPV | **0.77±0.04** | 0.70±0.01 | 0.68±0.03 | 0.67±0.02 | 0.69±0.02 | 0.61±0.02 |
| **External Dataset #2** | | | | | | |
| Accuracy | **0.68±0.06** | 0.53±0.05 | 0.58±0.06 | 0.55±0.07 | 0.63±0.04 | 0.47±0.04 |
| AUC | **0.79±0.02** | 0.72±0.04 | 0.67±0.04 | 0.65±0.03 | 0.74±0.04 | 0.76±0.04 |
| PPV | **0.39±0.04** | 0.29±0.02 | 0.30±0.02 | 0.28±0.03 | 0.34±0.02 | 0.28±0.02 |
| NPV | 0.93±0.03 | 0.92±0.02 | 0.87±0.01 | 0.85±0.02 | 0.91±0.02 | **0.96±0.01** |
| **External Dataset #3** | | | | | | |
| Accuracy | **0.68±0.06** | 0.58±0.08 | 0.62±0.03 | 0.61±0.03 | 0.66±0.04 | 0.64±0.05 |
| AUC | **0.72±0.03** | 0.57±0.02 | 0.62±0.03 | 0.60±0.03 | 0.67±0.03 | 0.44±0.05 |
| PPV | **0.46±0.08** | 0.32±0.05 | 0.36±0.03 | 0.35±0.02 | 0.41±0.04 | 0.20±0.09 |
| NPV | **0.83±0.02** | 0.75±0.02 | 0.79±0.03 | 0.78±0.02 | 0.81±0.01 | 0.71±0.02 |
| **External Dataset #4** | | | | | | |
| Accuracy | **0.71±0.03** | 0.63±0.06 | 0.63±0.07 | 0.62±0.07 | 0.67±0.07 | 0.68±0.05 |
| AUC | **0.82±0.02** | 0.72±0.02 | 0.65±0.04 | 0.64±0.03 | 0.73±0.03 | 0.63±0.09 |
| PPV | **0.38±0.02** | 0.31±0.04 | 0.30±0.04 | 0.29±0.05 | 0.33±0.06 | 0.29±0.08 |
| NPV | **0.93±0.02** | 0.89±0.02 | 0.87±0.02 | 0.86±0.02 | 0.88±0.03 | 0.83±0.02 |

Data are mean ± standard deviation. High means and low standard deviations indicated better performances.
Values in bold illustrated the best results.

**Table S5 Delong Test Results.**

| Datasets | D | E#1 | E#2 | E#3 | E#4 |
|---|---|---|---|---|---|
| HIC | 0.78 | 0.67* | 0.72* | 0.57* | 0.72* |
| HLR | 0.72 | 0.64* | 0.67* | 0.62* | 0.65* |
| Image Model | 0.70 | 0.62* | 0.65* | 0.60* | 0.64* |
| Clinical Model | 0.77 | 0.68* | 0.74* | 0.67 | 0.73* |
| Clinical FC | 0.77 | 0.52* | 0.76* | 0.44* | 0.63* |
| **MATE-Net** | **0.84** | **0.77** | **0.79** | **0.72** | **0.82** |

The DeLong test was applied to assess the AUCs. Average AUC values were presented, with * indicating statistical significance when compared to our MATE-Net.

D: Derivation dataset. E#1-E#4: External datasets #1-#4.

**Table S6 Performances of ablation studies**

| Baseline NIHSS | Tmax | Age | Accuracy | AUC | PPV | NPV |
|---|---|---|---|---|---|---|
| √ | √ | √ | **0.80±0.03** | **0.84±0.01** | **0.79±0.06** | **0.82±0.04** |
| × | √ | √ | 0.67±0.03 | 0.63±0.05 | 0.64±0.07 | 0.72±0.08 |
| √ | × | √ | 0.71±0.04 | 0.75±0.05 | 0.73±0.09 | 0.71±0.04 |
| √ | √ | × | 0.74±0.08 | 0.75±0.06 | 0.74±0.09 | 0.78±0.10 |
| × | × | √ | 0.55±0.05 | 0.52±0.05 | 0.51±0.07 | 0.58±0.04 |
| × | √ | × | 0.67±0.06 | 0.65±0.11 | 0.68±0.10 | 0.67±0.05 |

Values in bold illustrated the best results.

**Table S7 Correlations between C-SHAP and 90-day mRS, and correlations between other SHAP values and 90-day mRS.**

| Brain regions | Correlation coefficient (C-SHAP and 90-day mRS) | Correlation coefficient (SHAP values of baseline NIHSS and 90-day mRS) | Correlation coefficient (SHAP values of Tmax and 90-day mRS) | Correlation coefficient (SHAP values of age and 90-day mRS) |
|---|---|---|---|---|
| Posterior Temporal Lobe | 0.511* | 0.453* | 0.269* | 0.381* √ |
| **Superior Parietal Gyrus** | **0.511*** √ | **0.411*** | **0.302*** √ | **0.226*** |
| **Insula Posterior Long Gyrus** | **0.499*** √ | **0.470*** | **0.307*** | **0.203*** |
| Parahippocampal And Ambient Gyrus | 0.488* | 0.472* | 0.293* | 0.309* |
| Superior Temporal Gyrus Middle Part | 0.487* | 0.470* | 0.217* | 0.311* |
| Angular Gyrus | 0.477* | 0.421* | 0.263* | 0.364* |
| Superior Frontal Gyrus | 0.475* | 0.424* | 0.256* | 0.212* |
| **Precentral Gyrus** | **0.473*** √ | **0.460*** | **0.246*** | **0.258*** |
| Insula Anterior Long Gyrus | 0.472* | 0.464* √ | 0.268* √ | 0.228* |
| Supramarginal Gyrus | 0.472* | 0.476* | 0.199* | 0.388* √ |
| Postcentral Gyrus | 0.469* | 0.471* √ | 0.220* | 0.288* |
| Corpus Callosum | 0.468* | 0.434* | 0.090* | 0.358* |
| Pallidum | 0.467* | 0.395* | 0.245* | 0.308* |
| Middle And Inferior Temporal Gyrus | 0.464* | 0.464* | 0.151* | 0.339* √ |
| Posterior Cingulate Gyrus | 0.459* | 0.390* | 0.257* | 0.215* |
| Fusiform Gyrus | 0.459* | 0.474* | 0.296* | 0.254* |
| Nucleus Accumbens | 0.452* | 0.424* | 0.257* | 0.203* |
| Insula Posterior Short Gyrus | 0.446* | 0.462* | 0.254* | 0.264* |
| Middle Frontal Gyrus | 0.445* | 0.458* | 0.177* | 0.227* |
| Anterior Cingulate Gyrus | 0.443* | 0.439* | 0.198* | 0.252* |
| Insula Anterior Inferior Cortex | 0.439* | 0.419* | 0.248* | 0.162* |
| Putamen | 0.438* | 0.392* | 0.261* | 0.179* |
| Posterior Orbital Gyrus | 0.438* | 0.423* | 0.178* | 0.224* |
| Substantia Nigra | 0.435* | 0.441* | 0.170* | 0.185* |
| Inferior Frontal Gyrus | 0.427* | 0.475* | 0.209* | 0.130* |
| Straight Gyrus | 0.422* | 0.407* | 0.110* | 0.279* |
| Caudate Nucleus | 0.420* | 0.434* | 0.217* | 0.137* |
| Anterior Temporal Lobe Medial Part | 0.413* | 0.443* | 0.123* | 0.232* |
| Medial Orbital Gyrus | 0.411* | 0.453* | 0.170* | 0.165* |
| Insula Anterior Short Gyrus | 0.409* | 0.418* | 0.275* | 0.099* |
| Superior Temporal Gyrus Anterior Part | 0.405* | 0.438* | 0.156* | 0.064 |
| Pre-Subgenual Frontal Cortex | 0.404* | 0.410* | 0.086* | 0.198* |
| Subgenual Frontal Cortex | 0.399* | 0.388* | 0.024 | 0.224* |
| Subcallosal Area | 0.397* | 0.369* | 0.147* | 0.246* |
| Insula Middle Short Gyrus | 0.383* | 0.372* | 0.212* | 0.145* |
| Anterior Temporal Lobe Lateral Part | 0.380* | 0.456* | 0.132* | 0.112* |
| Anterior Orbital Gyrus | 0.323* | 0.403* | 0.114* | 0.123* |
| Lateral Orbital Gyrus | 0.258* | 0.362* | 0.147* | 0.124* |

* indicated the significance. √ expressed the consistently correlated brain regions. Regions in bold represented the selected brain regions.

**Table S8 Correlations between ischemic frequency and 90-day mRS**

| **Brain region** | **Correlation coefficient (Ischemic frequency and 90-day mRS)** | |
|---|---|---|
| Posterior Cingulate Gyrus | 0.225* | |
| **Pallidum** | **0.200*** | √ |
| Nucleus Accumbens | 0.196* | |
| Medial Orbital Gyrus | 0.183* | |
| Anterior Cingulate Gyrus | 0.176* | |
| **Precentral Gyrus** | **0.159*** | √ |
| Superior Frontal Gyrus | 0.154* | |
| Pre-Subgenual Frontal Cortex | 0.152* | |
| Subgenual Frontal Cortex | 0.151* | |
| Fusiform Gyrus | 0.149* | |
| Subcallosal Area | 0.148* | |
| Putamen | 0.145* | |
| Straight Gyrus | 0.143* | |
| Postcentral Gyrus | 0.138* | |
| Middle Frontal Gyrus | 0.137* | |
| Caudate Nucleus | 0.137* | |
| Corpus Callosum | 0.132* | |
| Parahippocampal And Ambient Gyrus | 0.127* | |
| Superior Parietal Gyrus | 0.126* | |
| Inferior Frontal Gyrus | 0.113* | |
| Insula Posterior Long Gyrus | 0.109* | |
| Anterior Orbital Gyrus | 0.104* | |
| Middle And Inferior Temporal Gyrus | 0.100* | |
| Posterior Orbital Gyrus | 0.100* | |
| Superior Temporal Gyrus Middle Part | 0.097* | |
| Posterior Temporal Lobe | 0.097* | |
| Insula Anterior Long Gyrus | 0.094* | |
| Insula Middle Short Gyrus | 0.094* | |
| Substantia Nigra | 0.092* | |
| Superior Temporal Gyrus Anterior Part | 0.092* | |
| Supramarginal Gyrus | 0.087* | |
| Lateral Orbital Gyrus | 0.086* | |
| Insula Anterior Short Gyrus | 0.083* | |
| Anterior Temporal Lobe Medial Part | 0.083* | |
| Insula Anterior Inferior Cortex | 0.082* | |
| Angular Gyrus | 0.078 | |
| Anterior Temporal Lobe Lateral Part | 0.076 | |
| Insula Posterior Short Gyrus | 0.074 | |

* indicated the significance. √ expressed the consistently correlated brain regions. Regions in bold represented the selected brain regions.

### Supplementary Figures

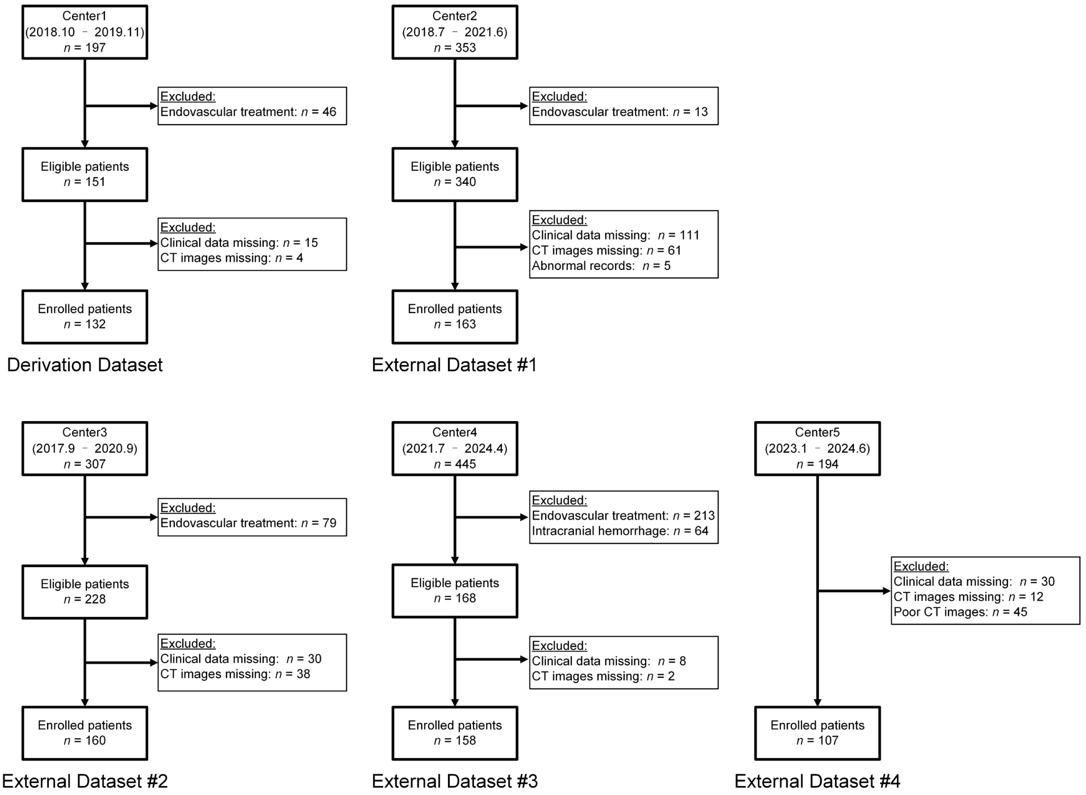


**Figure S1 Flow chart of patient enrollment.**

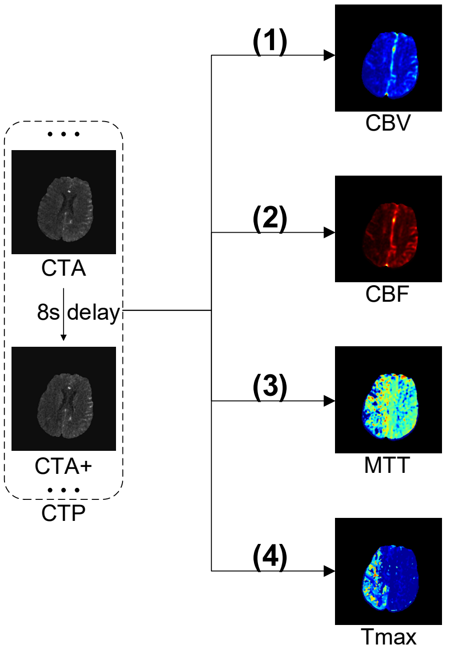


$$\text{(1)} \quad \mathrm{rCBV} = H \int c_t \, \mathrm{d}t$$

$$\text{(2)} \quad \mathrm{rCBF} = 60H \int c_a \, \mathrm{d}t[max_t(r(t))]$$

$$\text{(3)} \quad c_t(t) = r(t) \times c_a(t)$$

$$\text{(4)} \quad T_{max} = \arg max_t(r(t))$$

**Figure S2 Analysis of CT perfusion.** The meanings of the parameters are as follows: $H$ is a pre-defined constant, $c_t(t)$ is the perfusion image signal, $r(t)$ is tissue property, and $c_a(t)$ is arterial inflow function. CBV: cerebral blood volume. CBF: cerebral blood flow. MTT: mean transit time. Tmax: time-to-maximum.

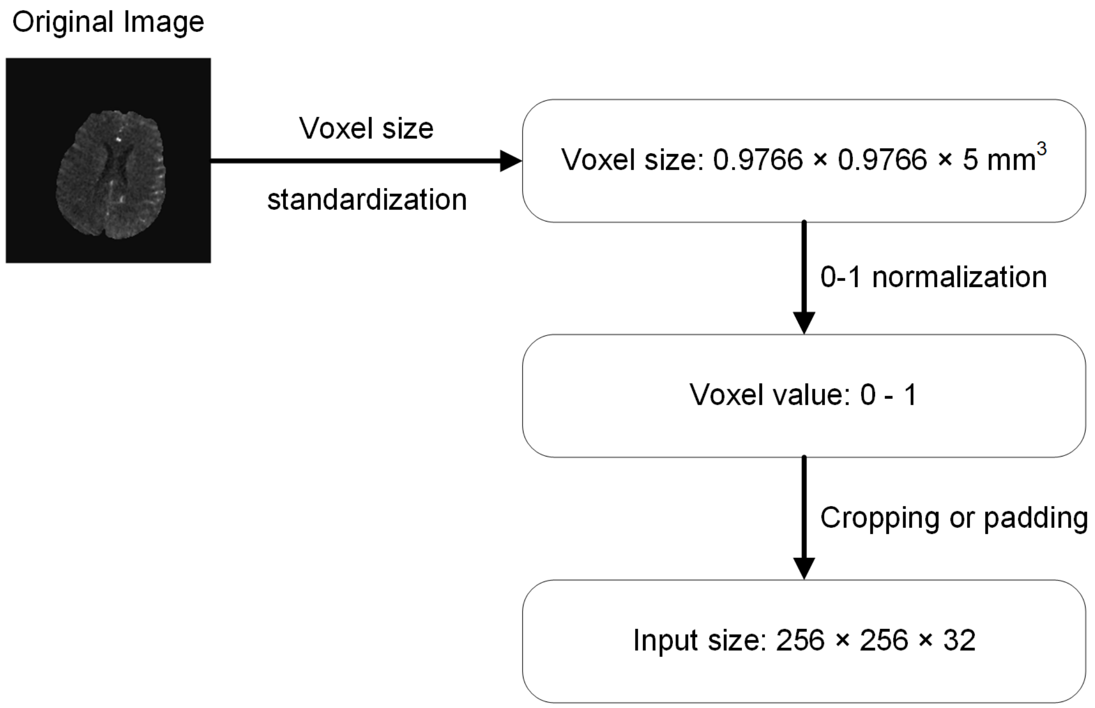


**Figure S3 Image preprocessing flowchart.**

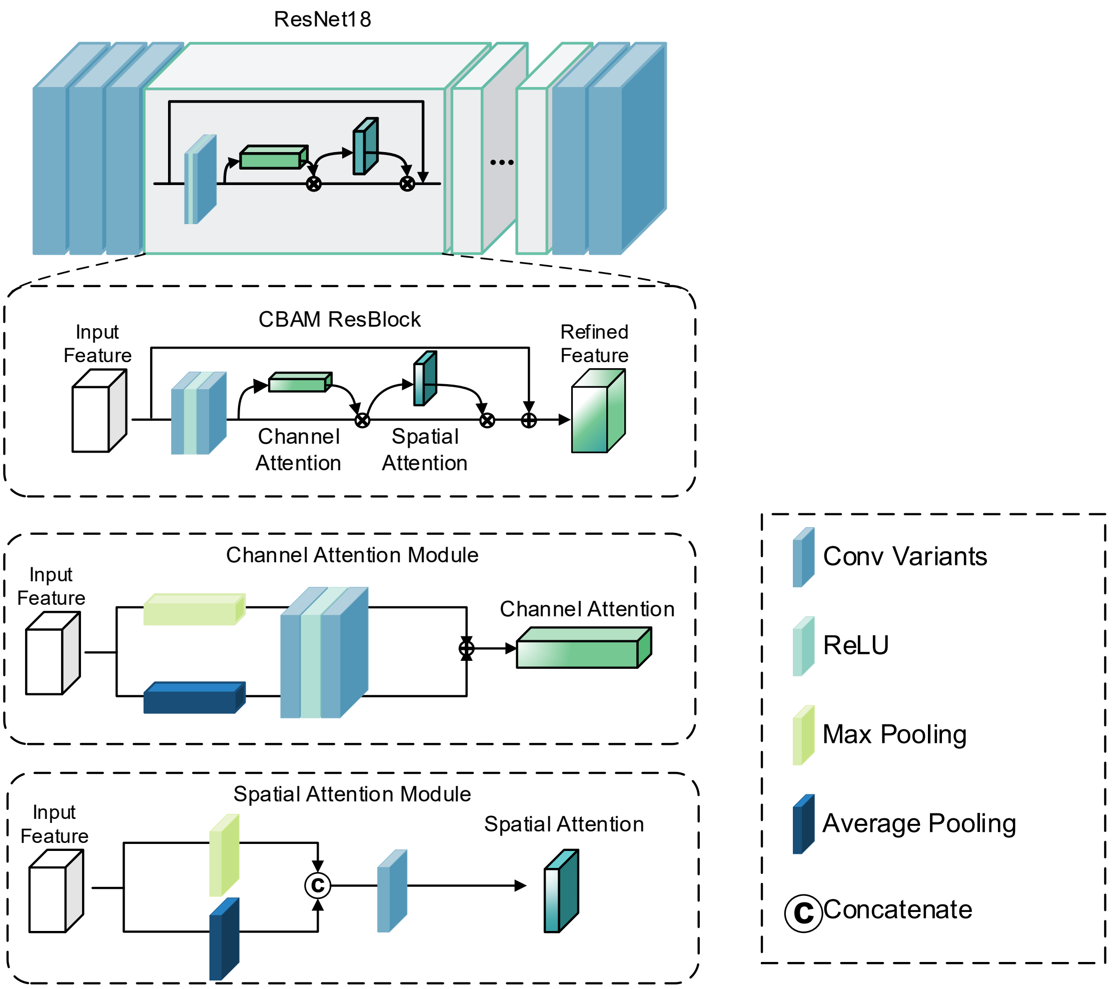


**Figure S4 Detailed structure of ResBlock and CBAM.** Detailed module parameters were seen in Supplementary Table I and Supplementary Table II. CBAM: Convolutional Block Attention Module.

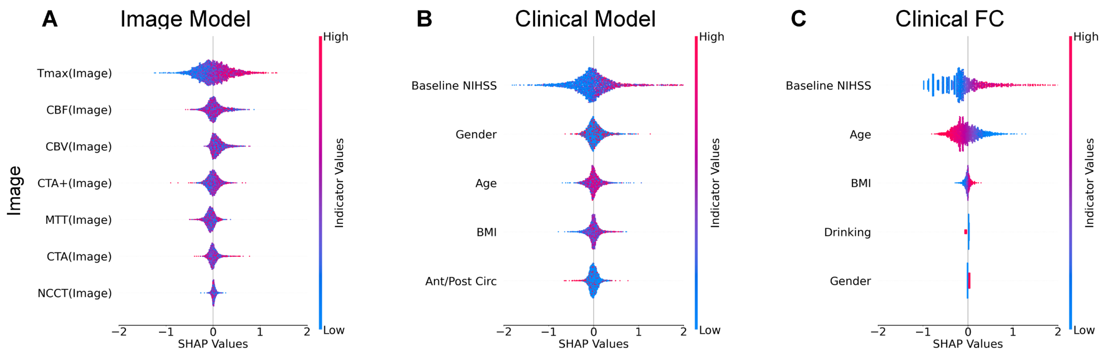


**Figure S5 Indicator ranking from other models.** SHAP values calculated for **(A)** Image Model (only CT images), **(B)** Clinical Model (only expanded clinical features) and **(C)** Clinical FC (Clinical Fully Connection model) are shown in decreasing order of mean absolute SHAP value. **(B)** and **(C)** 5 top indicators of SHAP value.

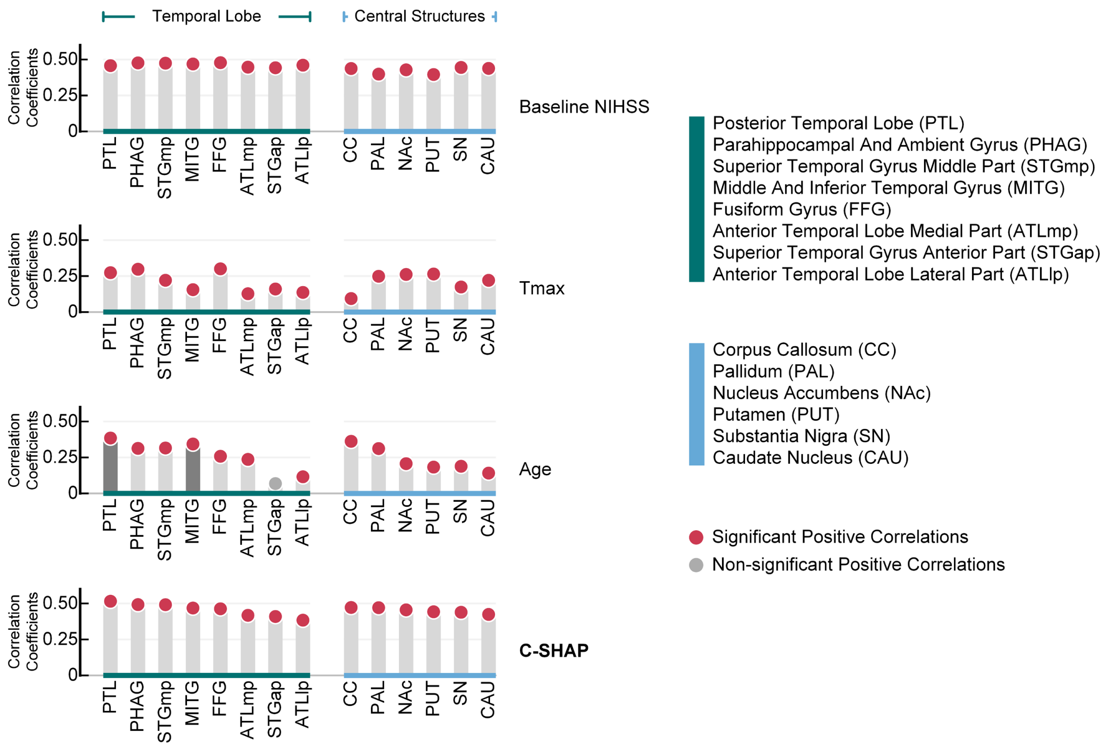


**Figure S6 Correlation coefficients for other sub anterior circulation brain regions.** C-SHAP obtained the highest correlation coefficients most compared with other separate indicators.
